# Two-Year Real World Clinical Outcomes after Intravascular Imaging Device Guided Percutaneous Coronary Intervention with Ultrathin-Strut Biodegradable-Polymer Sirolimus-Eluting Stent

**DOI:** 10.1101/2023.07.06.23292324

**Authors:** Sho Nakao, Takayuki Ishihara, Takuya Tsujimura, Yosuke Hata, Naoko Higashino, Masaya Kusuda, Toshiaki Mano

## Abstract

**Background:** Biodegradable-polymer sirolimus-eluting stents (BP-SES) have excellent technology, including ultrathin struts and nanocoating that suppresses metal ion elution, and have demonstrated improved results in numerous large clinical trials. However, many of these reports have not used intravascular imaging, and there is little clinical data on imaging-guided percutaneous coronary intervention (PCI) 1 year after the implantation when the polymer disappears. The current study investigated the clinical outcomes 2 years after imaging-guided PCI with BP-SES and durable-polymer everolimus-eluting stents (DP-EES).

**Methods:** We retrospectively analyzed 2455 patients who underwent successful PCI with BP-SES or DP-EES between September 2011 and March 2021, and compared 2-year clinical outcomes of BP-SES (459 patients) with DP-EES (1996 patients). The outcome measures were target lesion revascularization (TLR) and major adverse cardiac events (MACE), defined as a composite of cardiac death, myocardial infarction, target vessel revascularization, and stent thrombosis. Multivariate analysis using Cox proportional hazard model and inverse probability weighting (IPW) analysis based on the propensity score were used to evaluate the clinical outcomes.

**Results:** The 2-year cumulative incidences of TLR (BP-SES: 4.9% vs. DP-SES: 6.1%, p=0.304) and MACE (10.3% vs. 12.5%, p=0.159) were similar between the two groups. Multivariable and IPW analysis revealed the risks of TLR (p=0.388 and p=0.500) and MACE (p=0.139 and p=0.083) also had no significant difference. There was a significant interaction between none/mild and moderate/severe calcification with respect to MACE and TLR (adjusted p for interaction=0.036 and 0.029, respectively). The risk of MACE was significantly lower in BP-SES than in DP-EES in the lesions with none/mild calcification (adjusted hazard ratio [aHR]: 0.53; 95% confidence interval [CI]: 0.30-0.91), while it was similar in those with moderate/severe calcification (aHR: 0.95; 95% CI: 0.58-1.55).

**Conclusions:** Compared with DP-EES, BP-SES demonstrated durable 2-year clinical outcomes. However, BP-SES showed better clinical performance than DP-EES for lesions with none/mild calcification.

**Clinical Perspectives:** *What is Known:*

- Durable 1-year results of biodegradable-polymer sirolimus-eluting stents (BP-SES: Orsiro) for complex lesions such as acute coronary syndrome (ACS), small vessels or calcified lesions have been reported compared with contemporary thin-strut (81 mm) durable-polymer everolimus-eluting stents (DP-EESs, Xience).
- There are few reports comparing the long-term clinical outcomes of BP-SES and DP-EES in intravascular imaging device guided-percutaneous coronary intervention (PCI), although imaging-guided PCI has been reported to have better results than angiography-guided PCI.

*What the Study Adds:*

- BP-SES demonstrated comparable 2-year results after imaging-guided PCI with DP-EES.
- BP-SES demonstrated a lower risk of MACE in none/mild calcification and a similar risk in moderate/severe calcification as DP-EES.

## Introduction

A newly launched innovative third-generation drug-eluting stent (DES), biodegradable polymer sirolimus-eluting stent (BP-SES) (Orsiro BP-SES, Biotronik AG, Bulach, Switzerland), features a unique hybrid polymer laminate over ultrathin cobalt-chromium struts. The innermost layer of the ultrathin stent is arranged in a double helix pattern with 60 and 80 μm strut thickness for stent diameters less than or equal to 3 mm and more than 3 mm, respectively, designed to improve flexibility and deliverability. The middle proBIO™ layer confers a protective interface that guards against a reaction between the stent’s metal framework and the surrounding tissues. The outer BIOlute™ layer is composed of a bioabsorbable poly-L-lactic acid (PLLA) polymer containing an antiproliferative agent, sirolimus.^1^

Due to these excellent technologies, emerging evidences comparing ultrathin-strut versus thin-strut DESs indicated improved outcomes favoring ultrathin-strut DESs.^2, 3^ In addition, durable long-term results of BP-SES for complex lesions such as those of acute coronary syndrome (ACS) or small vessels, which are substantially encountered in real world clinical practice, have been reported compared with contemporary thin-strut (81 mm) durable-polymer everolimus-eluting stents (DP-EESs) (Xience, Abbott Vascular, Santa Clara, California).^4, 5^ Furthermore, although bench tests have shown that the radial force is weaker in thinner struts, clinical data reported that the BP-SES tended to be similar or better than the DP-EES in calcified lesions.^5–7^

However, many of these reports have not used intravascular imaging devices including intravascular ultrasound (IVUS) and optical coherence tomography (OCT), and there is little clinical data of imaging guided-percutaneous coronary intervention (PCI) after 1-2 years, when the polymer disappears.^8^ Imaging-guided PCI has been reported to have better results than angio-guided PCI, and may be useful for improving prognosis in complex lesions with high risk of restenosis and thrombosis.^9, 10^ PCI in Japan is characterized by a high frequency of imaging-guided PCI: however, to data, there are few reports comparing the long-term clinical outcomes of BP-SES and DP-EES in this procedure.^11^ Therefore, the current study investigated the 2-year clinical outcomes after imaging-guided PCI with BP-SES and DP-EES.

## Methods

### Study population

This was a single-center, retrospective, observational study. We retrospectively analyzed 2682 lesions in 2455 patients who underwent successful PCI with BP-SES (537 lesions in 459 patients) or DP-EES (2145 lesions in 1996 patients) between September 2011 and March 2021 and compared the 2-year clinical outcomes of BP-SES with DP-EES. Patients who underwent angiography-guided PCI or those who had out-of-hospital cardiopulmonary arrest were excluded from the study.

This study was performed in accordance with the Declaration of Helsinki and approved by the Ethics Committee of Kansai Rosai Hospital (approval no. 15D084g). Due to the retrospective and observational nature of the study, the need for written informed consent from patients was waived in accordance with the Ethical Guidelines for Medical and Health Research Involving Human Subjects in Japan. Instead, relevant information regarding the study was made available to the public and opportunities for individuals to refuse the inclusion of their data were ensured.

### Intervention procedure

Patients were eligible for inclusion if they had significant stenosis or occlusion on the initial coronary angiography and had undergone successful imaging-guided PCI with BP-SES or DP-EES. Intravascular imaging was performed using IVUS or OCT and PCI and post-PCI management, including antiplatelet therapy, were performed in a standard manner. Intravenous heparin (5000 IU), oral aspirin (200 mg), and prasugrel (20 mg) or clopidogrel (300 mg) were administered before PCI. After PCI, all patients received prasugrel (3.75 mg) or clopidogrel (75 mg) once daily in addition to aspirin (100 mg) for the optimal duration in accordance with relevant guidelines.^12, 13^

### Outcomes

The primary outcomes were the 2-year cumulative incidence of target lesion revascularization (TLR) and major adverse cardiac events (MACE), defined as a composite of cardiac death (CD), myocardial infarction (MI), target vessel revascularization (TVR), and stent thrombosis (ST). Secondary outcomes were other clinical outcomes, including all-cause death, CD, MI, TVR, non-target vessel revascularization (non-TVR), and definite ST.

### Definitions

Lesion calcification was assessed angiographically and classified according to a modified scheme of the American College of Cardiology (ACC) and American Heart Association (AHA) into: none or mild, moderate (visible on moving images during the heart cycle without contrast injection generally involving only 1 side of the arterial wall), and severe calcification (visible on still frame before contrast injection generally involving both sides of the arterial wall).^14^ ACS was defined as the presence of high-risk unstable angina (UAP), a non-ST elevation MI (NSTEMI), or an ST-elevation MI (STEMI). MI was diagnosed based on an increase in serum creatine phosphokinase, which was two-fold higher than the upper limit of the normal range, and had at least one of the following: symptoms of ischemia, new or presumed significant ST-segment-T wave (ST–T) changes or new left bundle branch block (LBBB), development of pathological Q waves in the electrocardiogram, imaging evidence of new loss of viable myocardium or new regional wall motion abnormalities, or identification of an intracoronary thrombus by angiography or autopsy.^15^ MI was defined as Type 1 to Type 3 or Type 4b based on the Third Universal Definition of Myocardial Infarction.^13^ TLR was defined as any clinically indicated repeat PCI of the target lesion or bypass surgery of the target vessel performed for restenosis or another complication of the target lesion.^15^ Revascularization was considered clinically indicated if angiography at follow-up showed a percent diameter stenosis of 50% or more and if one of the following was present: a positive history of recurrent angina pectoris, presumably related to the target vessel, objective signs of ischemia at rest or during an exercise test, presumably related to the target vessel, and abnormal results of any invasive functional diagnostic test.^16^ ST was defined according to the ARC definition.^12^

### Statistical analyses

All results are expressed as means ± standard deviations unless otherwise stated. Continuous variables with and without homogeneity of variance were analyzed using Student’s and Welch’s t-tests, respectively. Categorical variables were analyzed using Fisher’s exact test for 2 × 2 comparisons. For more than 2 × 2 comparisons, nominal and ordinal variables were analyzed using the chi-square and Mann–Whitney U tests, respectively. Clinical outcomes were evaluated using the Kaplan–Meier method and compared between BP-SES and DP-EES using the log-rank test. Additionally, in order to minimize inter-group differences of baseline characteristics, a multivariate Cox proportional hazard regression model was used to evaluate stent performance based on outcomes while adjusting for covariates including age, sex, ejection fraction, hypertension, dyslipidemia, diabetes mellitus, current smoking, chronic kidney disease, hemodialysis, chronic heart failure, stroke, atrial fibrillation, peripheral artery disease, type of ACS, angiotensin converting enzyme inhibitor (ACE-i)/angiotensin II receptor blocker (ARB) use, β-blocker use, mineralocorticoid receptor antagonist (MRA) use, statin use, ostial lesion, bifurcation, chronic total occlusion, moderate/severe calcification, ACC/AHA classification, in-stent restenosis, average stent size, total stent length, lesion location, number of stents, number of diseased vessels, and approach site. The results of the model were presented as hazard ratio (HR) and 95% confidence interval (CI). To confirm the robustness of the results, we performed an analysis using inverse probability weighting (IPW) based on the propensity score of the baseline characteristics. A logistic regression model was applied to predict the probability of clinical outcomes with the baseline covariates: age, sex, ejection fraction, hypertension, dyslipidemia, diabetes mellitus, current smoking, chronic kidney disease, hemodialysis, chronic heart failure, stroke, atrial fibrillation, peripheral artery disease, type of ACS, ACE-i/ARB use, β-blocker use, MRA use, statin use, ostial lesion, bifurcation, chronic total occlusion, moderate/severe calcification, ACC/AHA classification, in-stent restenosis, average stent size, total stent length, lesion location, number of stents, number of diseased vessels, and approach site. This was followed by the calculation of the HR of the stent type for outcomes with IPW based on the propensity score.

The interaction effects between the stent type and baseline patient and lesion characteristics were also assessed. HR and 95% CI were determined. A p-value less than 0.05 was considered statistically significant. All calculations were performed using IBM SPSS Statistics software version 28.0 J (IBM Corp., Armonk, NY, USA) and R software (version 4.0.3; R Foundation for Statistical Computing, Vienna, Austria; http://www.r-project.org/).

## Results

### Baseline characteristics

Patient, lesion, and procedural characteristics are summarized in **Table 1**. The proportion of male patients was higher, and the left ventricular ejection fraction (LVEF) was lower in the BP-SES group than in the DP-EES group. In terms of the coronary risk factors, hypertension was more common in the DP-EES group, while dyslipidemia and current smoking status were more common in the BP-SES group. BP-SES was more frequently used for ACS lesions Regarding the medication, MRAs and statins were more frequently prescribed in BP-SES group. Lesion complexities, including bifurcation and ACC/AHA classification Type B2/C lesions, were more severe in the BP-SES group, while ostial lesions and multiple vessel disease were more frequent in the DP-EES group. Moderate/severe calcification was similar between the two groups. Regarding procedural characteristics, the radial approach, pre-dilatation, and post-dilatation were more frequent, the post-dilatation balloon size was significantly larger, and the total stent length was significantly longer in the BP-SES group. The frequency of atherectomy device use was similar between the groups.

**Table 1.**
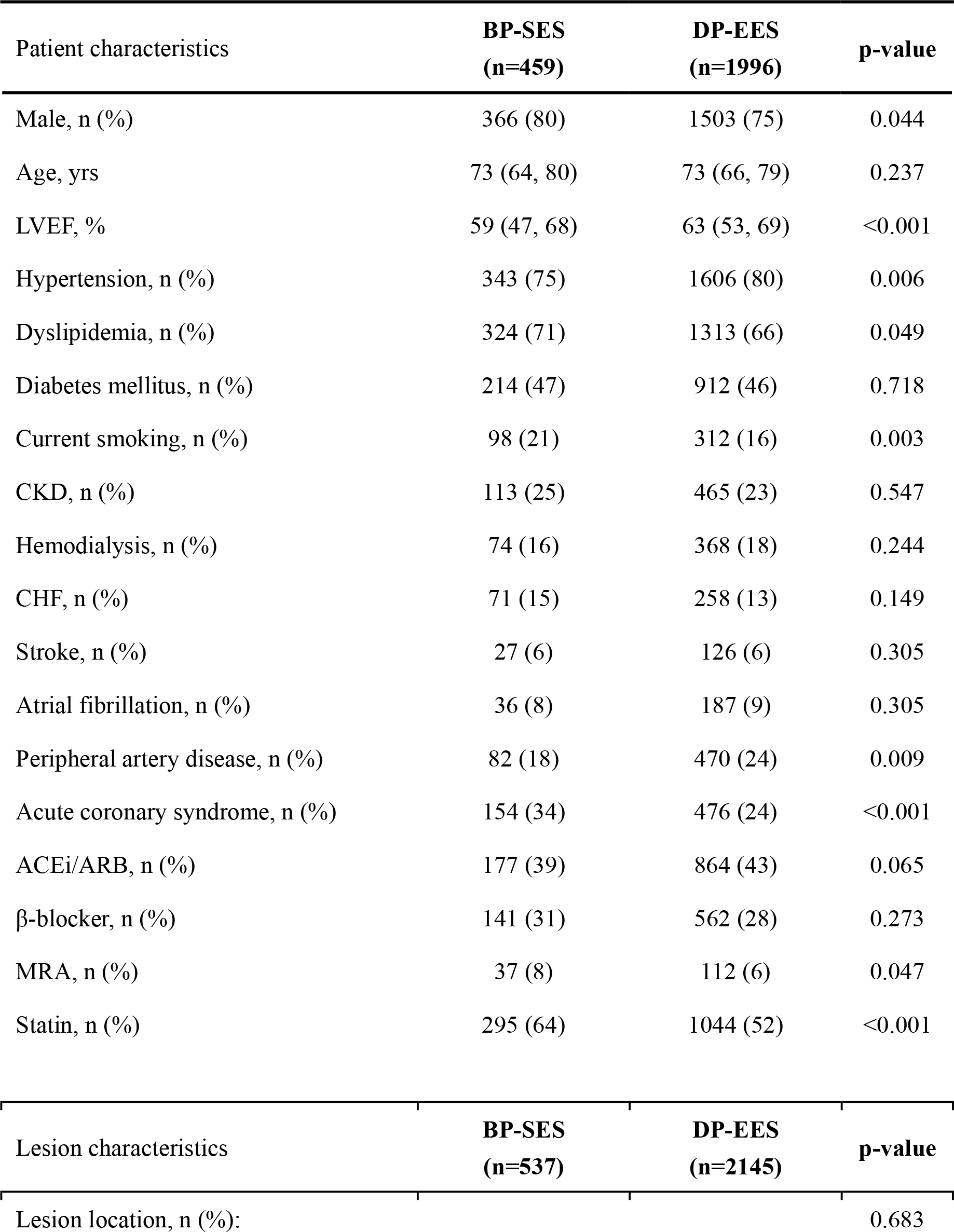
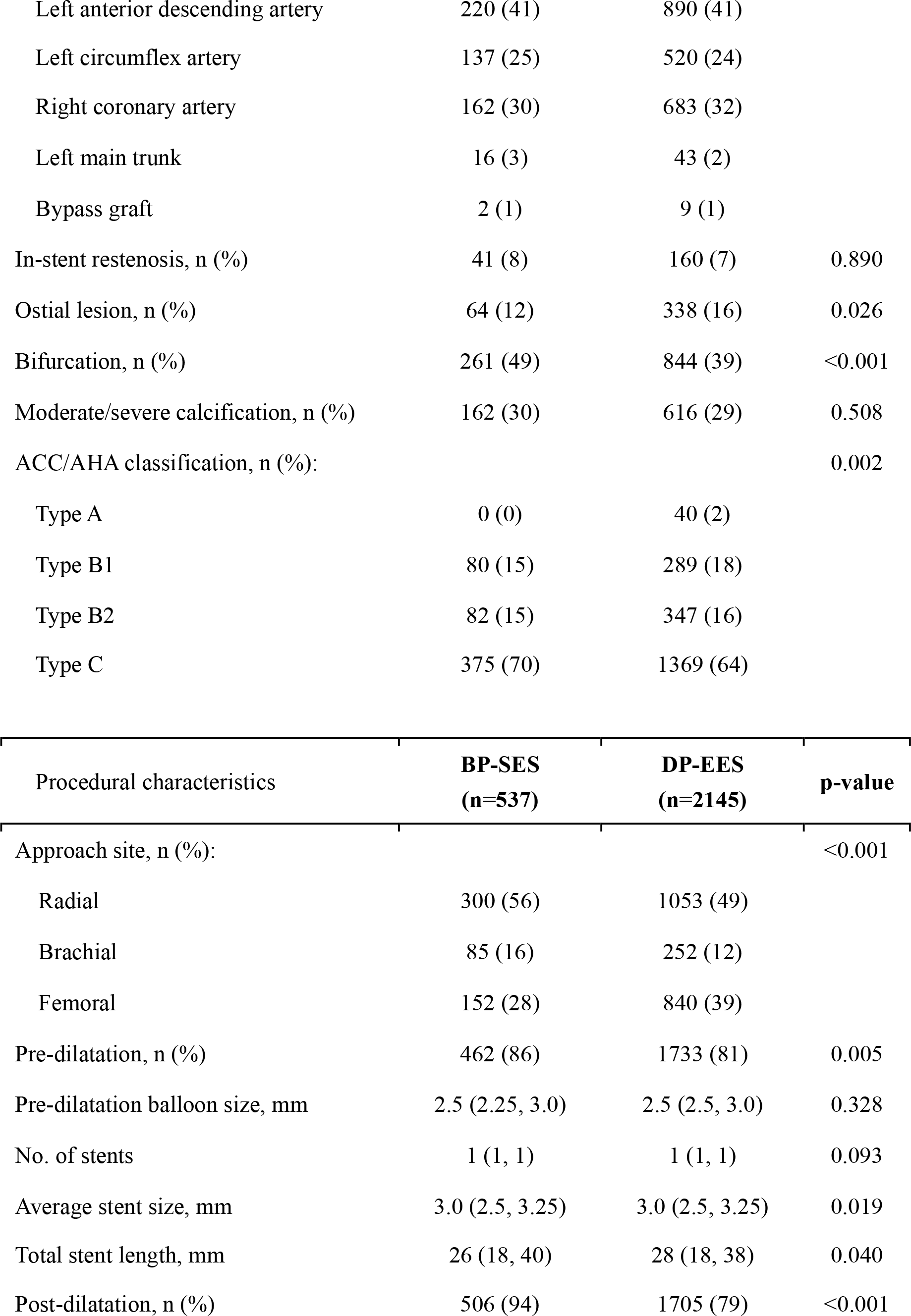
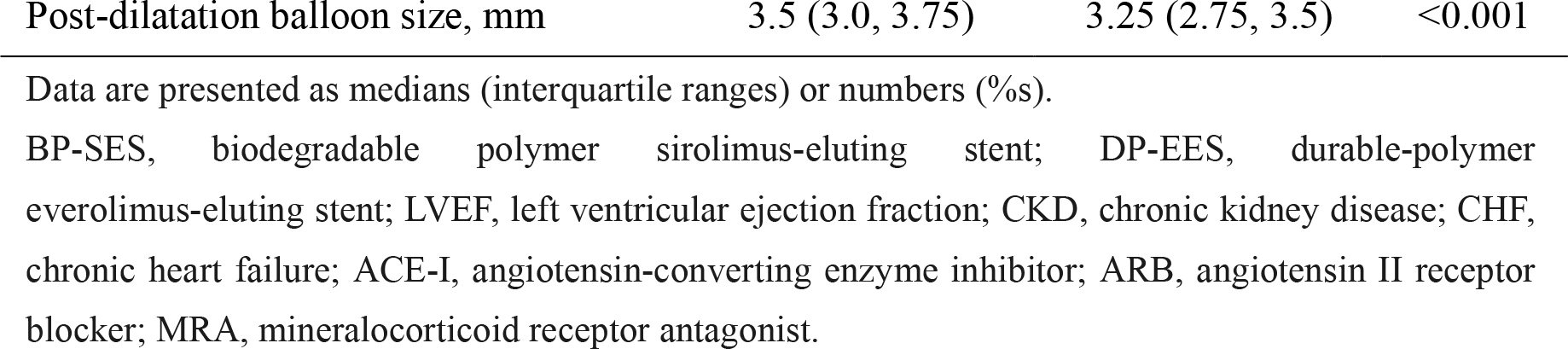
Patient, lesion, and procedural characteristics.

| Patient characteristics | **BP-SES**<br>**(n=459)** | **DP-EES**<br>**(n=1996)** | **p-value** |
| --- | --- | --- | --- |
| Male, n (%) | 366 (80) | 1503 (75) | 0.044 |
| Age, yrs | 73 (64, 80) | 73 (66, 79) | 0.237 |
| LVEF, % | 59 (47, 68) | 63 (53, 69) | <0.001 |
| Hypertension, n (%) | 343 (75) | 1606 (80) | 0.006 |
| Dyslipidemia, n (%) | 324 (71) | 1313 (66) | 0.049 |
| Diabetes mellitus, n (%) | 214 (47) | 912 (46) | 0.718 |
| Current smoking, n (%) | 98 (21) | 312 (16) | 0.003 |
| CKD, n (%) | 113 (25) | 465 (23) | 0.547 |
| Hemodialysis, n (%) | 74 (16) | 368 (18) | 0.244 |
| CHF, n (%) | 71 (15) | 258 (13) | 0.149 |
| Stroke, n (%) | 27 (6) | 126 (6) | 0.305 |
| Atrial fibrillation, n (%) | 36 (8) | 187 (9) | 0.305 |
| Peripheral artery disease, n (%) | 82 (18) | 470 (24) | 0.009 |
| Acute coronary syndrome, n (%) | 154 (34) | 476 (24) | <0.001 |
| ACEi/ARB, n (%) | 177 (39) | 864 (43) | 0.065 |
| β-blocker, n (%) | 141 (31) | 562 (28) | 0.273 |
| MRA, n (%) | 37 (8) | 112 (6) | 0.047 |
| Statin, n (%) | 295 (64) | 1044 (52) | <0.001 |

| Lesion characteristics | **BP-SES**<br>**(n=537)** | **DP-EES**<br>**(n=2145)** | **p-value** |
| --- | --- | --- | --- |
| Lesion location, n (%): |  |  | 0.683 |
| Left anterior descending artery | 220 (41) | 890 (41) |  |
| Left circumflex artery | 137 (25) | 520 (24) |  |
| Right coronary artery | 162 (30) | 683 (32) |  |
| Left main trunk | 16 (3) | 43 (2) |  |
| Bypass graft | 2 (1) | 9 (1) |  |
| In-stent restenosis, n (%) | 41 (8) | 160 (7) | 0.890 |
| Ostial lesion, n (%) | 64 (12) | 338 (16) | 0.026 |
| Bifurcation, n (%) | 261 (49) | 844 (39) | <0.001 |
| Moderate/severe calcification, n (%) | 162 (30) | 616 (29) | 0.508 |
| ACC/AHA classification, n (%): |  |  | 0.002 |
| Type A | 0 (0) | 40 (2) |  |
| Type B1 | 80 (15) | 289 (18) |  |
| Type B2 | 82 (15) | 347 (16) |  |
| Type C | 375 (70) | 1369 (64) |  |

| Procedural characteristics | **BP-SES**<br>**(n=537)** | **DP-EES**<br>**(n=2145)** | **p-value** |
| --- | --- | --- | --- |
| Approach site, n (%): |  |  | <0.001 |
| Radial | 300 (56) | 1053 (49) |  |
| Brachial | 85 (16) | 252 (12) |  |
| Femoral | 152 (28) | 840 (39) |  |
| Pre-dilatation, n (%) | 462 (86) | 1733 (81) | 0.005 |
| Pre-dilatation balloon size, mm | 2.5 (2.25, 3.0) | 2.5 (2.5, 3.0) | 0.328 |
| No. of stents | 1 (1, 1) | 1 (1, 1) | 0.093 |
| Average stent size, mm | 3.0 (2.5, 3.25) | 3.0 (2.5, 3.25) | 0.019 |
| Total stent length, mm | 26 (18, 40) | 28 (18, 38) | 0.040 |
| Post-dilatation, n (%) | 506 (94) | 1705 (79) | <0.001 |
| Post-dilatation balloon size, mm | 3.5 (3.0, 3.75) | 3.25 (2.75, 3.5) | <0.001 |

Data are presented as medians (interquartile ranges) or numbers (%s).
BP-SES, biodegradable polymer sirolimus-eluting stent; DP-EES, durable-polymer everolimus-eluting stent; LVEF, left ventricular ejection fraction; CKD, chronic kidney disease; CHF, chronic heart failure; ACE-I, angiotensin-converting enzyme inhibitor; ARB, angiotensin II receptor blocker; MRA, mineralocorticoid receptor antagonist.

### Clinical outcomes

**Table 2** and **Figure 1** show the cumulative incidence of each outcome and its Kaplan–Meier curve. Regarding primary outcomes, the cumulative incidences of TLR and MACE were similar between the two groups. The cumulative incidences of all-cause death, CD, MI, TVR, non-TVR, and ST were not significantly different between the two groups. Furthermore, after adjusting for covariates using a multivariate Cox proportional hazard regression model, the cumulative incidences of TLR and MACE, as well as those of other clinical outcomes, were not significantly different between the two groups (**Table 3**). The IPW analysis consistently showed a similar risk of TLR and MACE in the two groups.

**Table 2:** Cumulative incidence of each clinical outcome.

| **Outcome** | **BP-SES** | **DP-EES** | **p-value** |
| --- | --- | --- | --- |
| All cause death, n (%) |  |  | 0.968 |
| Number of events | 42 | 185 |  |
| 2-year event rate | 10.2±0.7 | 10.0±1.5 |  |
| MACE, n (%) |  |  | 0.159 |
| Number of events | 41 | 224 |  |
| 2-year event rate | 10.3±1.5 | 12.5±0.8 |  |
| Cardiac death, n (%) |  |  | 0.682 |
| Number of events | 14 | 54 |  |
| 2-year event rate | 3.5±0.9 | 2.9±0.4 |  |
| MI, n (%) |  |  | 0.116 |
| Number of events | 2 | 26 |  |
| 2-year event rate | 0.5±0.4 | 1.5±0.3 |  |
| TLR, n (%) |  |  | 0.304 |
| Number of events | 22 | 112 |  |
| 2-year event rate | 4.9±1.0 | 6.1±0.6 |  |
| TVR, n (%) |  |  | 0.249 |
| Number of events | 37 | 181 |  |
| 2-year event rate | 8.1±1.3 | 9.8±0.7 |  |
| Non-TVR, n (%) |  |  | 0.216 |
| Number of events | 49 | 237 |  |
| 2-year event rate | 10.7±1.5 | 13.0±0.8 |  |
| ST, n (%) |  |  | 0.361 |
| Number of events | 1 | 10 |  |
| 2-year event rate | 0.2±0.2 | 0.5±0.2 |  |

Data are presented as means ± standard deviations and percentages.
BP-SES, biodegradable polymer sirolimus-eluting stent; DP-EES, durable-polymer everolimus-eluting stent; TLR, target lesion revascularization; MACE, major adverse cardiovascular events; MI, myocardial infarction; TVR, target vessel revascularization; non-TVR, non-target vessel revascularization; ST, stent thrombosis.

**Table 3:** Cumulative incidence of each clinical outcome after adjusting for covariates by a multivariate Cox proportional hazard regression model and IPW.

|  | Crude |  | Multivariate |  | IPW |  |
| --- | --- | --- | --- | --- | --- | --- |
|  | HR | p-value | HR | p-value | HR | p-value |
| TLR | 0.787 [0.498-1.243] | 0.304 | 0.807 [0.497-1.312] | 0.388 | 0.838 [0.501-1.401] | 0.500 |
| MACE | 0.788 [0.565-1.099] | 0.159 | 0.770 [0.544-1.089] | 0.139 | 0.722 [0.499-1.044] | 0.083 |
| All cause death | 0.993 [0.710-1.388] | 0.968 | 0.859 [0.602-1.224] | 0.400 | 0.936 [0.641-1.366] | 0.731 |
| Cardiac death | 1.131 [0.628-2.035] | 0.682 | 0.670 [0.346-1.300] | 0.236 | 0.767 [0.404-1.458] | 0.419 |
| MI | 0.334 [0.079-1.408] | 0.116 | 0.420 [0.096-1.831] | 0.248 | 0.447 [0.104-1.914] | 0.278 |
| TVR | 0.813 [0.571-1.157] | 0.249 | 0.851 [0.589-1.230] | 0.391 | 0.872 [0.570-1.241] | 0.384 |
| Non-TVR | 0.824 [0.606-1.121] | 0.216 | 0.853 [0.621-1.172] | 0.328 | 0.872 [0.624-1.219] | 0.423 |
| ST | 0.396 [0.051-3.096] | 0.361 | 0.464 [0.044-4.839] | 0.521 | 0.619 [0.008-4.817] | 0.647 |

IPW, inverse probability weighting; HR, hazard ratio; TLR, target lesion revascularization; MACE, major adverse cardiovascular events; MI, myocardial infarction; TVR, target vessel revascularization; non-TVR, non-target vessel revascularization; ST, stent thrombosis.

**Fig. 1.**
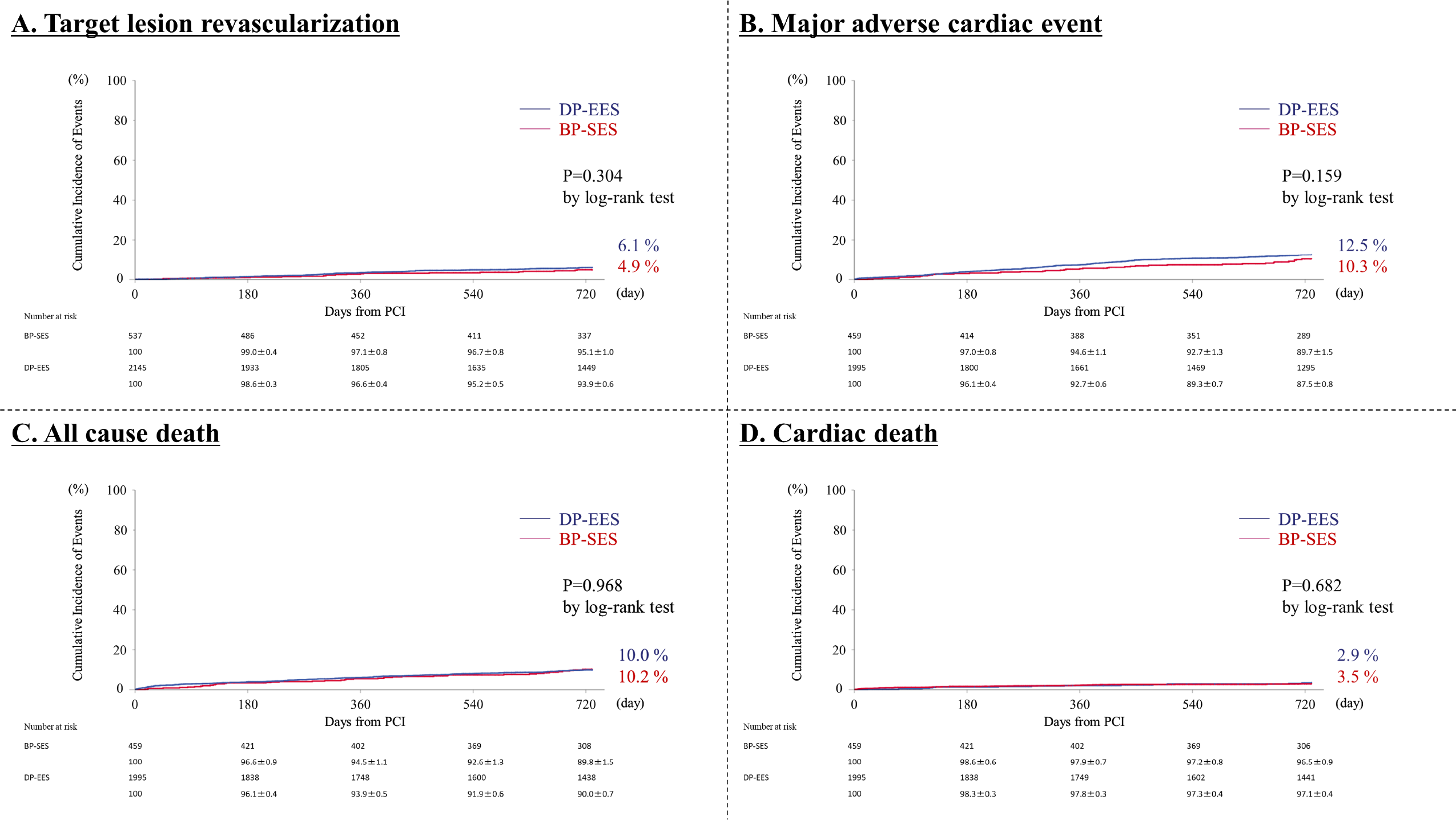
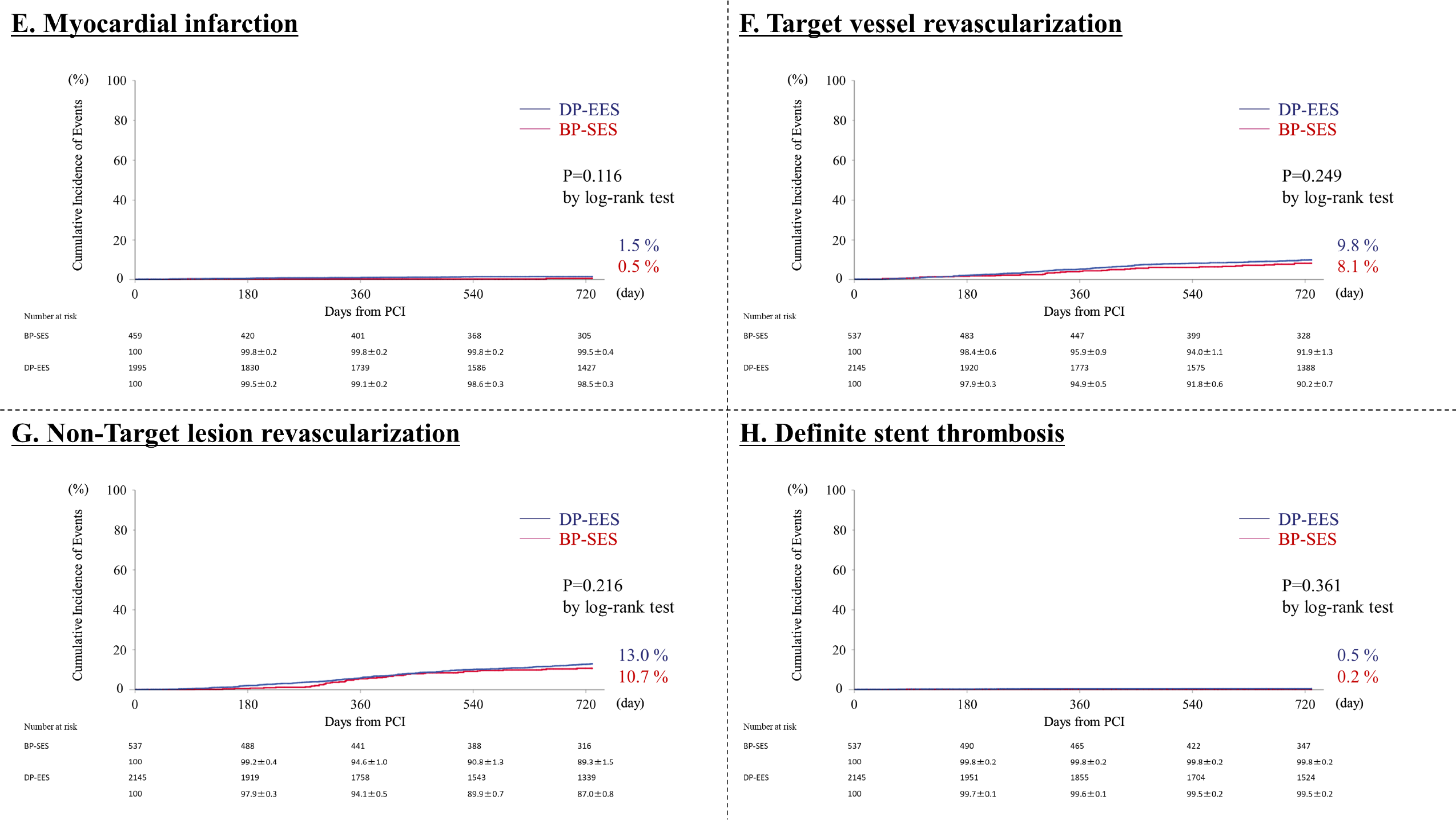
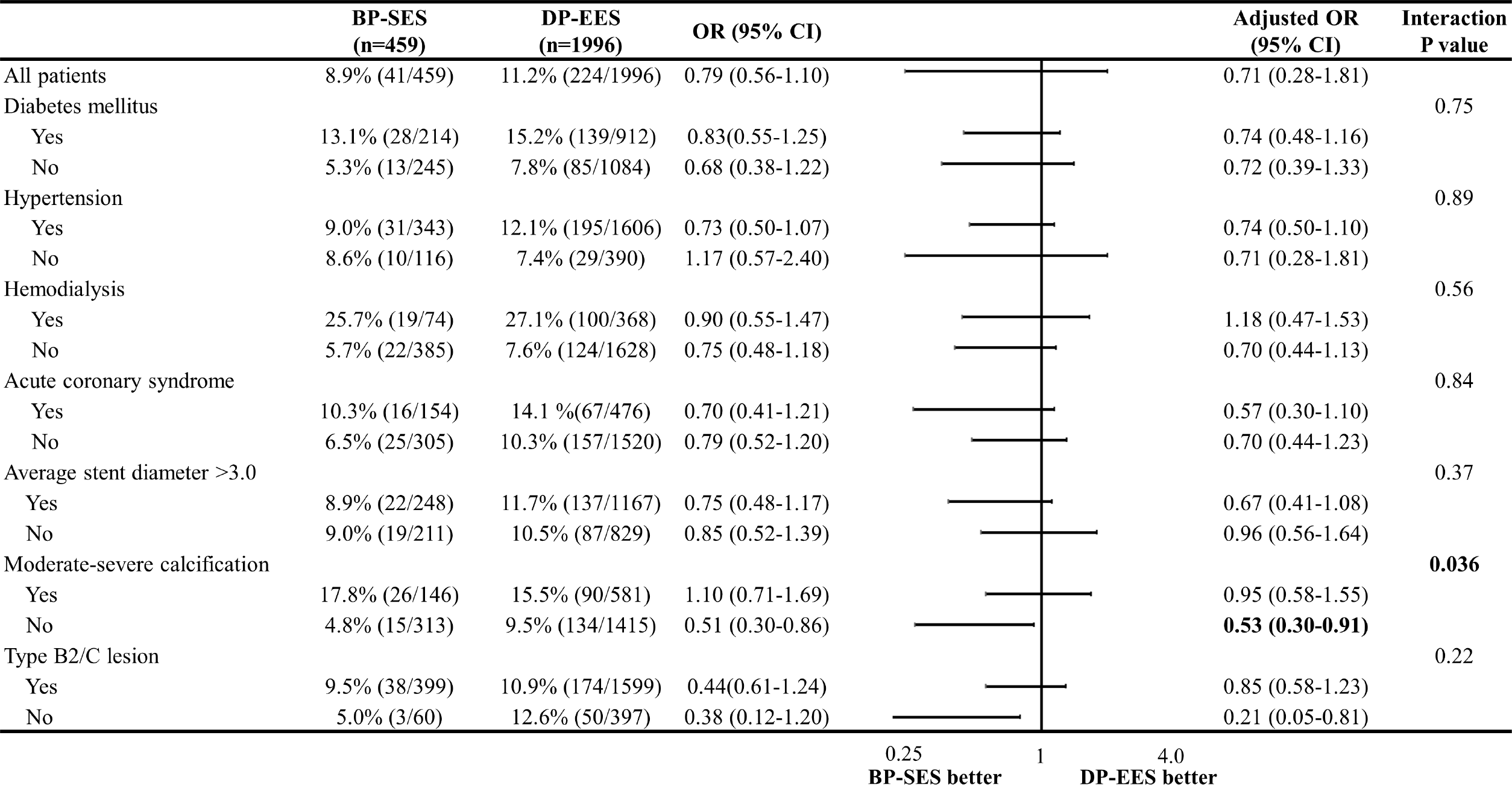
Cumulative incidence of 2-year clinical outcomes. A: Target lesion revascularization, 4.9%; BP-SES, 6.1%; DP-EES, (p=0.304). B: Major adverse cardiac events,10.3%; BP-SES, 12.5%; DP-EES, (p=0.159). C: All-cause death:10.2%, BP-SES and 10.0%, DP-EES (p=0.968). D: Cardiac death,3.5%; BP-SES, 2.9%; DP-EES, (p=0.682). E: Myocardial infarction,0.5%; BP-SES, 1.5%; DP-EES, (p=0.116). F: Target vessel revascularization,8.1%; BP-SES, 9.8%; DP-EES, (p=0.249). G: Non-target vessel revascularization,10.7%; BP-SES, 13.0%; DP-EES, (p=0.216). H: Definite stent thrombosis,0.2%; BP-SES, 0.5%; DP-EES, (p=0.361). BP-SES, biodegradable polymer sirolimus-eluting stent; DP-EES, durable-polymer everolimus-eluting stent.

Regarding the interaction effects between the stent type and baseline patient and lesion characteristics, there were significant interactions between none/mild and moderate/severe calcification, and the risk of MACEs was significantly lower in the BP-DES group (**Figure 2**). **Figure 3** shows the Kaplan–Meier curves of MACE stratified by none/mild calcification and moderate/severe calcification between the two groups. BP-SES demonstrated a lower risk of MACE in none/mild calcification and a similar risk in moderate/severe calcification as DP-EES.

**Fig. 2.**
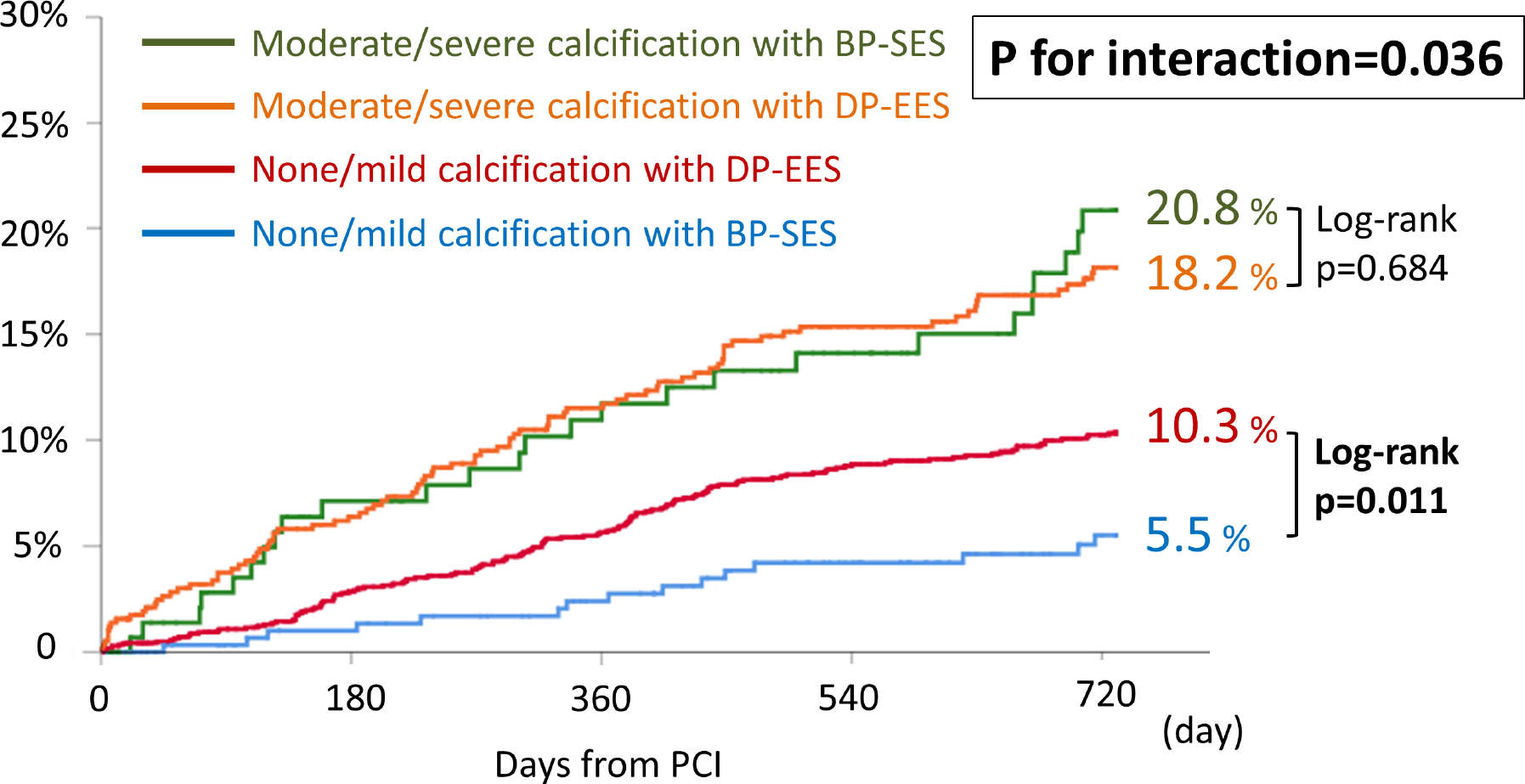
Interaction effect of BP-SES versus DP-EES on MACE in the baseline characteristics. There were significant interactions between none/mild and moderate/severe calcification, and the risks were significantly lower in the BP-DES group with regard to MACE (p for interaction=0.036). BP-SES, biodegradable polymer sirolimus-eluting stent; DP-EES, durable-polymer everolimus-eluting stent; MACE. major adverse clinical outcomes.

**Fig. 3.**
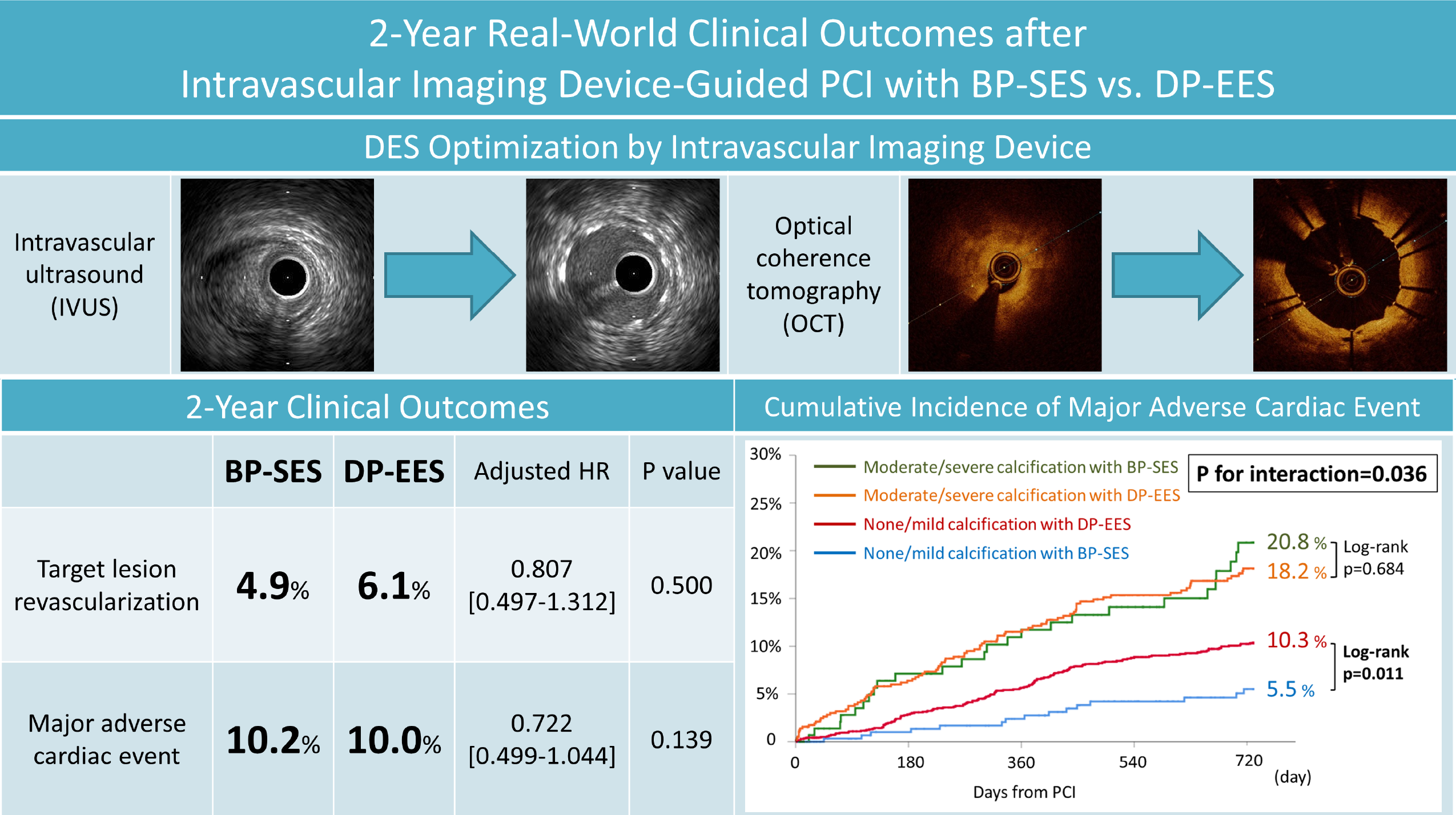
Cumulative incidence of 2-year MACE stratified by none/mild and moderate/severe calcification between BP-SES and DP-EES. BP-SES demonstrated a lower risk of MACE in none/mild calcification and a similar risk in moderate/severe calcification as DP-EES. BP-SES, biodegradable polymer sirolimus-eluting stent; DP-EES, durable-polymer everolimus-eluting stent; MACE. major adverse clinical outcomes.

## Discussion

The results of our retrospective analyses of 2455 patients who underwent successful imaging-guided PCI with BP-SES or DP-EES at our hospital demonstrated that the cumulative incidence of TLR and MACE was similar between the two groups in both the multivariate and IPW analysis. Interaction analysis revealed that none/mild calcification showed better results in the BP-SES group; however, no interaction was observed in moderate/severe calcification between the two groups. To the best of our knowledge, this is the first study to systematically demonstrate the real world 2-year clinical performance of imaging-guided PCI with BP-SES and DP-EES.

### The 2-year clinical performance of imaging-guided PCI with BP-SES

Although the current study revealed that patient background (age: 73 years vs. 70 years, diabetes mellites: 47% vs. 39%, chronic kidney disease: 25% vs. 16%, ACS: 34% vs. 15%) and lesion background (bifurcation: 49% vs. 32%, moderate/severe calcification: 30% vs. 20%) were both complicated, the cumulative incidence of 1-year MACE was 5.4%, which was similar to the previously reported data that 1-year target lesion failure after imaging-guided PCI with BP-SES was 6% and durable results were also observed at 2 years.^8^ However, at 2 years after the PCI with BP-SES, when the polymer disappeared, the results were still comparable to DP-EES. This demonstrates that strut design differences among DESs, including stent strut thickness or polymer coating, have little impact on clinical outcomes, even 2 years after implantation with intravascular imaging guidance. Further follow-up is necessary to prove the advantage of ultrathin or proBIO nanocoating, which reduce thrombogenicity and promote endothelialization after biodegradable polymer degradation. The pre-specified 3-year follow-up data of randomly assigned patients in the CASTLE trial are expected to help elucidate whether imaging-guided DES implantation has a long-term impact on clinical outcomes following PCI.

### BP-SES for complex lesions

Although it seems reasonable to consider that patients with ACS, who are in a relatively high thrombotic state, and patients with small-vessel disease in which stents occupy a relatively greater amount for the vessel luminal diameter, are the ideal candidates for treatment with ultrathin-strut DESs, the current study revealed no interactions in ACS lesions or small vessels, for which previous studies reported the efficacy of BP-SES over DP-EES.^5^ This would be due to reductions in procedure-related suboptimal DES implantation thanks to the imaging-guided PCI compared with angiography-guided PCI.^17, 18^ Intravascular imaging devices help determine the appropriate stent diameter, stent length, and stent placement position based on quantitative evaluations of the lumen, vascular diameter, and plaque volume. They also enable detection of inadequate stent expansion or stent edge dissection, which may help predict stent thrombosis.^19–22^ In addition to quantitative evaluation, qualitative evaluations of plaque morphology and distribution are also available, which predict distal embolism, side branch occlusion, or necessity for plaque modification such as atherectomy or lithotripsy before stent placement.^23^ These characteristics make intravascular imaging guidance more effective in complex lesions such as small vessels and ACS lesions.

### BP-SES for calcified lesions

In this study, none/mild calcification showed better results with BP-SES with imaging-guided PCI; however, no interaction was observed in moderate/severe calcification. This result is consistent with a previous report of angiography-guided PCI.^7^ In none/mild calcification, technological advances such as ultrathin-struts with biodegradable-polymer proBIO coatings on a double helix stent design may have led to better results even under the conditions of optimal stent placement with imaging-guided PCI. However, this difference was not observed for moderate/severe calcification. While the effectiveness of imaging-guided PCI in calcified lesions has been reported, it has been pathologically demonstrated that calcified lesions pose a greater risk of stent thrombosis due to delayed healing, indicated by uncoverage, and restenosis due to excessive neointimal proliferation resulting from severe medial tear leading.^18^ Therefore, the advantages of ultrathin stents are diminished in calcified lesions, and the potential of the stent platform becomes smaller. Furthermore, no interactions were observed with the use of an atherectomy device in this study; however, as atherectomy was performed in only 7% of the population, the effectiveness of BP-SES may have been underestimated in lesions with sufficient atherectomy. In situations with adequate lesion preparation, the advantages of stent platform characteristics are maximized, and the effectiveness of BP-SES can be demonstrated.

### Limitations

This study had several limitations. First, it was a single-center retrospective observational study, and there was heterogeneity between the two groups as noted in the baseline demographic and procedural variables. However, the sample size was large, and we matched the baseline characteristics with multivariate Cox regression and IPW analysis based on propensity score. Second, although we newly reported the 2-year clinical outcomes after imaging-guided PCI, the efficacy of the nanocoating may become more obvious after 2 years when the polymer is completely absorbed. Longer follow-up results from the CASTLE study are expected. Third, calcification characteristics, including the thickness, depth, and number of calcified nodules, were not considered. In this study, calcification was evaluated using contrast-guided imaging according to previous reports. Owing to the limited use of OCT, it was difficult to quantitatively assess calcification parameters. Therefore, further studies using OCT are required. Fourth, intravascular lithotripsy was unavailable in Japan during the study period and therefore, it was not used. It is now available, and further data accumulation is expected in the future.

## Conclusion

The BP-SES demonstrated durable 2-year clinical outcomes compared with the DP-EES. However, BP-SES showed better clinical performance than DP-EES for lesions with none/mild calcification.

## Data Availability

All of the authors have full access to all the data in the study and takes responsibility for its integrity and the data analysis.

## Nonstandard Abbreviations and Acronyms

ACS: acute coronary syndrome
BP-SES: biodegradable-polymer sirolimus-eluting stent
DP-EES: durable-polymer everolimus-eluting stent
IPW: inverse probability weighting
IVUS: intravascular ultrasound
LVEF: left ventricular ejection fraction
MACE: major adverse cardiac events
MI: myocardial infarction
PCI: percutaneous coronary intervention
ST: stent thrombosis
TLR: target lesion revascularization
TVR: target vessel revascularization

## Acknowledgments

We wish to thank Ms. Saori Kashu for her expertise in data aggregation.

